# Decreased Mitochondrial Respiration in Peripheral Mononuclear Blood Cells in Children and Adolescents with Obesity and Type 2 Diabetes Mellitus

**DOI:** 10.1101/2025.09.30.25336982

**Authors:** Marlene Rechtsteiner, Samiya Al-Robaiy, Hans Zischka, Susanne Kröber, Andreas Simm, Paulo J. Oliveira, Eugénia Carvalho, Susann Weihrauch-Blüher

## Abstract

**Background:** The prevalence of childhood obesity continues to rise worldwide. Obesity leads to major health risks already early in life, including insulin resistance (IR) and type 2 diabetes mellitus (T2DM). The pathogenesis of these conditions includes mitochondrial alterations, however, data on mitochondrial health in pediatric populations are scarce to date.

**Methods:** The study evaluated mitochondrial respiration in peripheral blood mononuclear cells (PBMCs) from children and adolescents (6–18 years) with obesity with different stages of IR or T2DM, respectively. Participants were stratified according to pubertal stage and metabolic status. Mitochondrial health was determined by key parameters of mitochondrial respiration (ATP production, coupling efficiency, proton leak), and the Bioenergetic Health Index (BHI) served as an integrative marker of mitochondrial performance.

**Results:** A total of 162 participants were included. The Bioenergetic Health Index (BHI) was significantly lower in postpubertal T2DM adolescents compared to postpubertal IR participants, despite of equal pubertal stage and BMI (p=0,0126). The mitochondrial respiration rates and BHI between prepubertal and peripubertal children with obesity and IR showed no statistical differences. However, GlycoATP was correlated to higher insulin and HbA1c levels in children with obesity and IR. In line, the T2DM group demonstrated significantly reduced coupling efficiency (p = 0.003), reduced mitoATP and elevated glycoATP production, indicating impaired mitochondrial efficiency.

**Conclusion:** These data suggest for the first time, that progression of IR towards manifest T2DM in children with obesity leads to impaired mitochondrial function. Thus, mitochondrial alterations in PBMCs may detect early metabolic impairment in young people with obesity and altered glucose metabolism.

**Graphical Abstract:** 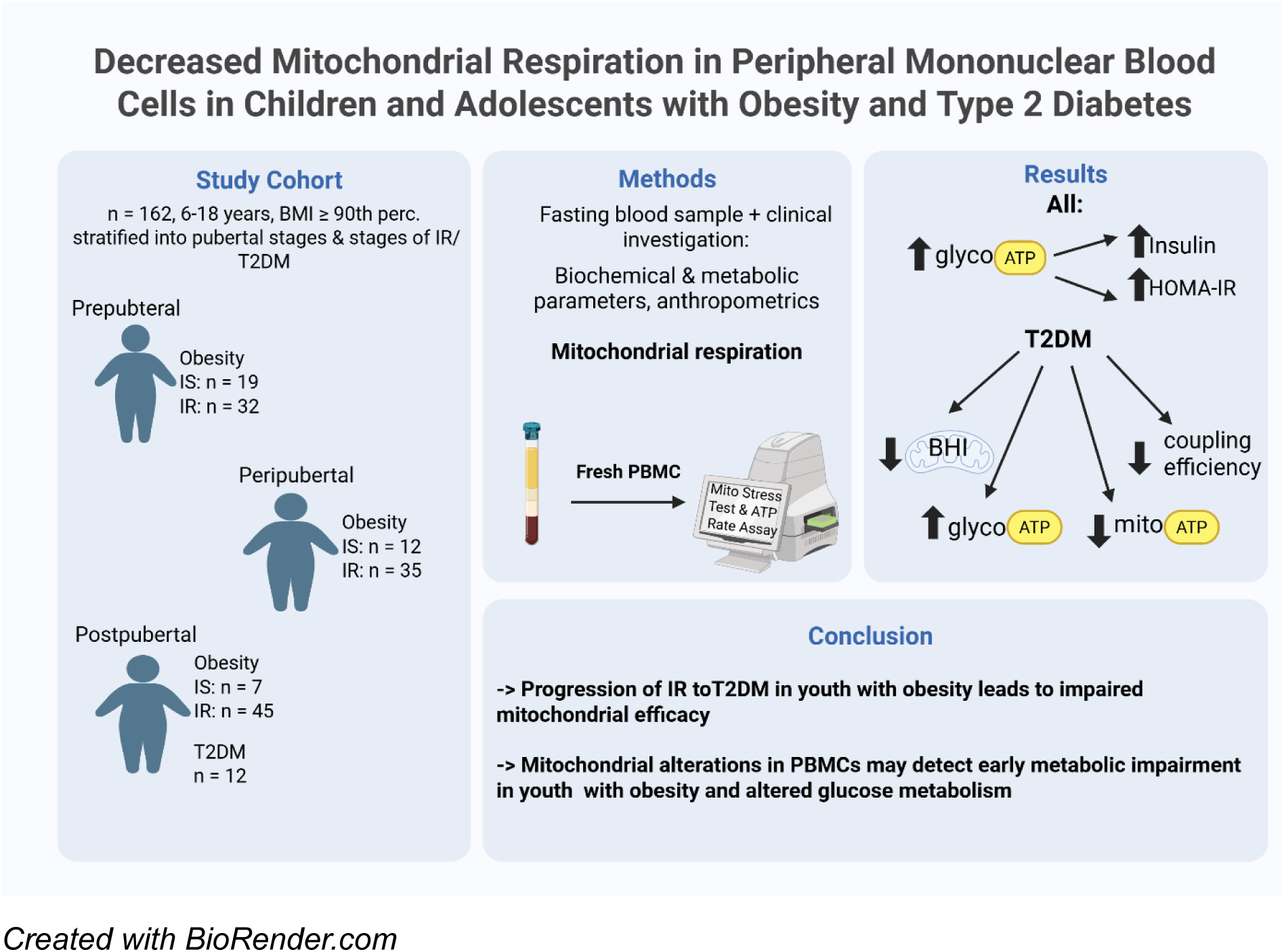

**Highlights:**

- GlycoATP is correlated to higher insulin and HbA1c levels in children with obesity and
- Insulin resistance (IR)
- Adolescents with T2DM have significantly reduced coupling efficiency, reduced mito-
- ATP and elevated glycoATP production, indicating impaired mitochondrial efficiency
- Data suggest for the first time, that progression of IR towards manifest T2DM in children with obesity leads to impaired mitochondrial function
- Mitochondrial alterations in PBMCs may detect early metabolic impairment in young
- people with obesity and altered glucose metabolism.

## 1. Introduction

Childhood obesity has emerged as one of the most pressing public health challenges of the 21st century, with its prevalence increasing worldwide. According to the World Health Organization (WHO), the prevalence of overweight in Europe in the age group 5-9 years increased to 29,5 % and for obesity to 11,6 % within the last 30 years [1]. Childhood obesity causes multiple serious physical and psychological problems, and it serves as a primary risk factor for Metabolic Syndrome and Type 2 Diabetes Mellitus (T2DM), as well as cardiovascular disease and other conditions [2, 3]. Even in early childhood, obesity and associated morbidities can lead to long-term consequences in health, persisting into adulthood [4]. This underscores the importance of early detection and intervention.

The incidence of youth-onset T2DM has increased worldwide by 30 % over the past two decades in line with the increasing obesity prevalence and unfavorable lifestyle factors such as high consumption of sugar-sweetened beverages and an delayed diagnosis because of its long subclinical course and lacking awareness among patients and caregivers [5]. Nonetheless, it still remains a rare diagnosis in adolescents, and number of adolescent patients with manifest T2DM who are treated in specialized centers are still small [6].

Mitochondria are essential organelles responsible for energy production through oxidative phosphorylation, playing also other central roles in cellular metabolism. Mitochondrial dysfunction is one of the key elements responsible for metabolic disease development, including insulin resistance and subsequent development of the Metabolic Syndrome (MetS) as well as subclinical inflammation [7].

In the context of obesity, calorie excess and body fat accumulation can lead to mitochondrial dysfunction [8, 9]. The mitochondrial dysfunctional state produces impaired oxidative phosphorylation while generating higher reactive oxygen species (ROS) levels that lead to oxidative stress and subsequent damage of cellular components and worsening of metabolic disorders [10].

Evidence shows that T2DM development is associated with mitochondrial dysfunction which causes insulin resistance as one of the main downstream complications. The dysfunction of mitochondrial oxidative phosphorylation and other important mitochondrial pathways disrupt insulin signaling pathways, which results in reduced glucose uptake and utilization. The impaired function relates to changes in mitochondrial activity and dynamics that ultimately decrease the activity of the mitochondrial respiratory chain, reducing ATP generation and increased ROS production, while other mitochondrial functions such as calcium regulation is also affected [11].

Mitochondrial dysfunction-associated oxidative stress together with systemic inflammation constitute key elements of the Metabolic Syndrome. 19The relationship between mitochondrial impairment and metabolic disturbance shows that mitochondrial integrity is crucial in preventing and treating metabolic disorders. The investigation of these mechanisms supports the development of innovative therapeutic strategies and improved early metabolic dysfunction detection methods [12].

Studies on obesity commonly measure mitochondrial dysfunction in adipocytes together with skeletal cells [13]. Peripheral blood mononuclear cells (PBMCs) including lymphocytes and monocytes provide researchers with a practical approach to assess mitochondrial function through minimally invasive clinical assessments, especially important in pediatric populations. Therefore, the accessibility of PBMCs through standard blood testing makes them an excellent choice for pediatric research applications [14]. In fact, research has demonstrated that mitochondrial function in PBMCs correlates with skeletal muscle respiration and other known clinical parameters [15, 16].

Comparative analyses between PBMCs and other cell types have further validated the use of PBMCs in pediatric populations. In a study investigating mitochondrial function across different human cell types, PBMCs were utilized to assess age-related declines in mitochondrial activity, underscoring their applicability in pediatric research [17].

Alterations in PBMC metabolism have been observed in adult populations with obesity and relating metabolic conditions, such as T2DM, indicating the potential of PBMCs as indicators for systemic metabolic health [18, 19]. However, to the best of our knowledge there are not studies that have investigated metabolic health in PBMC cells in children and adolescents. The aim of this study is thus to identify differences in mitochondrial respiration in fresh PBMCs between children and adolescents with obesity and different states of insulin resistance as well as adolescents with T2DM. In addition, we aimed to analyze if mitochondrial respiration differs depending on the severity of the metabolic impairment as well as depending on pubertal stage.

## 2. Materials and Methods

### 2.1 Study design

This study is part of an EU-funded multicenter project PAS GRAS „*Derisking metabolic, environmental and behavioral determinants of obesity in children, adolescents and young adults”* which investigates individual risk factors for the development of obesity and its consequential diseases from early childhood to adulthood.

Written consent was obtained by the Ethical Commission of University medicine Halle, Germany, (# 2023-238). The study was performed in accordance with the declaration of Helsinki and based on the STROBE criteria. Written informed consent was obtained from the parents or legal guardians of all participants. Children aged 6 to 16 provided assent to participate, while participants over the age of 16 also provided informed consent.

In this study two groups of participants were included:

Group 1: Children adolescents aged 6-18 years with obesity (BMI ≥90^th^ percentile according to German reference values [20].

Group 2: Adolescents with obesity and manifest T2DM according to WHO guidelines [21], aged 6-19 years with BMI ≥90, percentile according to German reference values [20].

Exclusion criteria were syndromal obesity, pregnancy, and chronic diseases that prevent study participation.

Participants were stratified into four groups based on pubertal development and the presence of IR or T2DM: prepubertal (Tanner stage 1), peripubertal (Tanner stages 2–3), postpubertal (Tanner stages 4–5), and individuals with manifest T2DM.

For the different analyses, participants were further stratified based on insulin sensitivity. Insulin resistance (IR) was defined using the homeostasis model assessment of insulin resistance (HOMA-IR) with a cutoff at the 90th percentile for age and sex. Three subgroups were formed: (1) insulin sensitive children (HOMA-IR < 90^th^ percentile) (IS), (2) insulin-resistant (IR), and (3) participants with T2DM.

### 2.2 Anthropometric measurements and biochemical parameters

Measurements of body weight and lengths of all participants were performed in light underwear following standardized procedures. Body mass index (BMI) was calculated by the formula: weight in kilograms divided by the square of the height in meters. BMI values were transformed to standard deviation scores (BMI-SDS) based on German reference values [20]. A cut off ≥1.28 SDS (90th centile) is applied to classify overweight, a cut off ≥ 1.88 SDS (97th centile) to classify obesity, and a cut-off ≥ 2.58 SDS (99.5th centile) to classify extreme obesity [22].

Waist circumstance was measured using a flexible band following the standard procedures. Waist to height ratio (WHtR) was calculated by dividing the waist circumstance in cm with the height in cm. WHtR serves as a validated measure for visceral obesity, and a WHtR ≥ 0,5 is associated with higher cardiometabolic risk [23, 24]. Pubertal staging was assessed by trained clinicians according to standard Tanner criteria [25].

A fasting blood sample was obtained, and the following parameters were analyzed: glucose, insulin, HbA1c, serum uric acid, (sUA), Alanine Aminotransferase (ALAT), Aspartate Aminotransferase (ASAT), total cholesterol, HDL-Cholsesterol, triglycerides at the laboratory of the University clinic Halle (Saale), Germany by standardized procedures. Homeostasis Model Assessment of Insulin Resistance (HOMA-IR) was calculated by multiplying fasting glucose [mmol/l] with fasting insulin [mU/l] divided by 22,5. HOMA-IR is used as an index for severity of insulin resistance [26]. Age adapted percentiles were used to categorize the HOMA-IR into normal and disturbed insulin resistance [27]. HOMA-IR below the 90^th^ percentile was considered normal insulin sensitivity, ≥90^th^ percentile insulin resistance, and ≥97^th^ percentile severe insulin resistance.

### 2.3 Isolation of PBMCs

PBMCs were isolated from fasting, venous blood samples using the Leucosep preparation tubes from Greiner Bio-One. The 10 ml Leucosep tube was filled with 3 ml room temperature (RT) Pancoll human 1.077 g/ml density gradient medium (PAN Biotech) and spun for 1 min with 1000x g with brakes on. Seven ml whole blood was layered on top of the porous barrier and the tube was spun at 1000x g for 10 min at RT with brakes switched off. The enriched cell fraction (buffy coat) was harvested using Pasteur pipettes. The harvested cells were washed with 10 ml phosphate-buffered saline (PBS) and spun at RT for 10 min with 1000x g, brakes on. To remove any residual erythrocytes, the cells were treated with 3 ml erythrocyte lysis buffer (c-c-pro mbH). After washing the cells again with 13 ml PBS, they were counted and viability checked with trypan blue staining and a Neubauer improved counting chamber. The PBMCs where instantly used for Extracellular Flux Analysis, as described below.

## 2.4 Measurement of bioenergetics in PBMCs

Measurements were performed with PBMC activity, Seahorse XF96 Analyser XF96 Analyzer (RRID:SCR_019545) using the wave software (Agilent, Santa Clara, USA). The 96-well plate was coated with poly-D-lysine (Gibco^TM^) the day before the assay and stored at 4°C upside down. On the day of the assay, PBMCs are suspended in Seahorse XF RPMI Medium (pH 7,4) containing 10 mM glucose, 1 mM sodium pyruvate and 2 mM glutamine (Agilent, Santa Clara, USA). An amount of 300,000 cells per well was plated in 80 µl assay medium. The plate was centrifuged in one direction at 50x g, RT, brakes off for 5 minutes and in the other direction at 80x g, RT, brakes off for 5 minutes. Each well was carefully filled up to a total volume of 175 µl. The plate was incubated at 37 °C without CO_2_ for 60 minutes before running the assay.

The Agilent Seahorse XF Cell Mito Stress Test and the Agilent Seahorse XF Real-Time ATP Rate Assay (Agilent, Santa Clara, USA) were applied on respectively one half of the plate according to the instructions to measure the oxygen consumption rates [28]. The sensor cartridges, hydrated the day prior to the assay, were filled with the compound solutions of the mitochondrial respiration modulators. For the Cell Mito Stress Test, port A was filled with 20 µl oligomycin (final concentration 2 µM), port B with 22 µl Carbonyl cyanide-4 (trifluoromethoxy) phenylhydrazone (FCCP) (final concentration 2 µM) and port C with 25 µl rotenone and antimycin A (final concentration 0,5 µM). For the Real-Time ATP Rate Assay, port A was filled with 20 µl oligomycin (final concentration 2 µM), port B with 22 µl rotenone and antimycin A (final concentration 0,5 µM) and port C with 25 µl assay medium. After the calibration of the sensor cartridge, the cell plate was launched to the Seahorse Analyzer and analyzed with 4 repetitions of each cycle.

The parameters of the Mito Stress Test were calculated from normalized raw data according to the manufacturers manual [29]. Following the respiratory data obtained, the Bioenergetic Health Index (BHI) was calculated as described by Chacko et al. [30]. The BHI integrates multiple parameters derived from the mitochondrial stress test, including ATP-linked respiration, proton leak, spare respiratory capacity, and non-mitochondrial respiration. The BHI was calculated using the following formula:

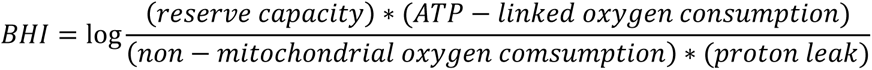

This composite index provides a sensitive and quantitative measure of mitochondrial bioenergetic health under physiological and stress conditions. A higher index indicates better mitochondrial health [31].

The parameters of the ATP Rate Assay were calculated using the Oxygen Consumption Rates (OCR) and Extracellular Acidification Rate (ECAR) in the Agilent Seahorse Wave Software. Mitochondrial ATP production was calculated as: *mitoATP production rate (pmol ATP/min) = OCR_ATP_ (pmol O_2_/min) * 2 (pmol O/pmol O_2_) * P/O (pmol ATP/pmol O)*. Glycolic ATP Production rate was calculated as: *glycoATP Production Rate (pmol ATP/min) = glycoPER (Pmol H^+^/min*). Due to high variations in absolute ATP values in between measurements, relative ratios of mitoATP and glycoATP were analyzed [32].

### 2.5 Statistical analyses

Descriptive statistics were used to summarize demographic, anthropometric, biochemical, and mitochondrial parameters. Continuous variables are presented as means with standard deviation (mean ± SD) or as medians with interquartile ranges (Q1, Q3), depending on data distribution. Categorical variables are presented as counts and percentages.

For group comparisons across metabolic subgroups, defined by Tanner-Stages, HOMA-IR percentiles and the presence of type 2 diabetes, one-way analysis of variance (ANOVA) was used to assess differences in mitochondrial function and clinical parameters. Where appropriate, post-hoc analyses were applied to further investigate group-wise differences.

In all children without manifest T2DM Spearman correlation analysis were performed between mitochondrial respiration and ATP production data and parameters associated with IR and T2DM. A p-value < 0.05 was considered statistically significant for all analyses.

All statistical analyses were conducted using IBM SPSS Statistics Version 28.0 and GraphPad Prism software (version 10.2.0).

## 3. Results

A total of 162 participants were stratified into four groups according to pubertal stage: prepubertal (Tanner stage 1, n = 51), peripubertal (Tanner stages 2-3, n = 47), postpubertal (Tanner stages 4-5, n = 52), and a postpubertal group with T2DM (n = 12). Mean age in the prepubertal group was significantly lower than in the postpubertal / T2DM group (10.0±1.7 vs. 15.1±2.4 years; p < 0.001).

The sex distribution did not differ significantly between groups (p = 0.265). BMI-SDS increased significantly during puberty (p = 0.006), peaking in the postpubertal group (2.91 ± 0.68). HOMA-IR scores demonstrated a significant increase across the groups (p < 0.001) with the highest values recorded in participants with T2DM (10.4 ± 8.3). The percentage of participants with insulin resistance (IR) (HOMA-IR ≥ 90th percentile) compared to insulin sensitive (IS) participants rose from 62.7% in the prepubertal group to 86.5% in the postpubertal group. Detailed group characteristics are summarized in Table 1.

**Table 1:** Descriptive statistics from study population. P-values for continuous variables from one-way ANOVA, and for categorical variables from Chi-square test.

| Variables | Prepubertal<br>Tanner 1<br>N = 51 | Peripubertal<br>Tanner 2 & 3<br>N = 47 | Postpubertal<br>Tanner 4 & 5<br>N = 52 | T2DM<br>Tanner 4 & 5<br>N= 12 | P -<br>Value |
| --- | --- | --- | --- | --- | --- |
| **Metformin** |  |  |  |  |  |
| Treatment | 6 (11.8 %) | 17 (36.2 %) | 41 (78.8 %) | 12 (100 %) |  |
| No Treatment | 45 (88.2 %) | 30 (63.8 %) | 11 (21.2 %) | 0 (0 %) |  |
| **Sex** |  |  |  |  |  |
| Female | 19 (37.3 %) | 21 (44.7 %) | 27 (51.9 %) | 3 (25 %) | 0.260 |
| Male | 32 (62.7 %) | 26 (55.3 %) | 25 (48.1 %) | 9 (75%) |  |
| **Age [years]** |  |  |  |  |  |
| Mean ± SD | 10.0 ± 1.7 | 12.3 ± 1.7 | 15.2 ± 1.7 | 15.1 ± 2.4 | <0.001 |
| Median (Q1, Q3) | 10.2 (9.1, 11.1) | 12.5 (11.3, 13.5) | 13.5 (10.8, 18.8) | 15.1 (13.2, 16.9) |  |
| **BMI-SDS** |  |  |  |  |  |
| Mean ± SD | 2.56 ± 0.46 | 2.57 ± 0.53 | 2.91 ± 0.68 | 2.53 ± 0.77 | 0.006 |
| Median (Q1, Q3) | 2.55 (2.24; 2.94) | 2.62 (2.23, 2.88) | 2.93 (2.62, 3.26) | 2.65 (2.03, 2.93) |  |
| **WHtR** |  |  |  |  |  |
| Mean ± SD | 0.57 ± 0.06 | 0.58 ± 0.07 | 0.61 ± 0.08 | 0.59 ± 0.07 | 0.054 |
| Median (Q1, Q3) | 0.59 (0.53, 0.62) | 0.58 (0.54, 0.64) | 0.62 (0.59, 0.66) | 0.59 (0.53, 0.65) |  |
| **HOMA-IR** |  |  |  |  |  |
| Mean ± SD | 3.8 ± 2.1 | 5.5 ± 3.2 | 6.2 ± 3.0 | 10.4 ± 8.3 | <0.001 |
| Median (Q1, Q3) | 3.1 (2.4, 3.5) | 5.1 (3.3, 7.2) | 6.5 (5.4, 8.8) | 6.5 (4.2, 17.6) |  |
| **Insulin-Resistance** |  |  |  |  |  |
| (HOMA-IR ≥ 90.<br>Percentile) | 32 (62.7 %) | 35 (74.5 %) | 45 (86.5 %) | | |
| **Fasting glucose** |  |  |  |  |  |
| **[mmol/l]** |  |  |  |  |  |
| Mean ± SD | 4.85 ± 0.37 | 5.06 ± 0.45 | 4.93 ± 0.37 | 6.66 ± 1.58 | <0.001 |
| Median (Q1, Q3) | 4.80 (4.62, 5.14) | 5.03 (4.76, 5.30) | 4.95 (4.76, 5.27) | 6.56 (5.53, 6.96) |  |
| **Fasting Insulin** |  |  |  |  |  |
| **[mU/l]** |  |  |  |  |  |
| Mean ± SD | 17.32 ± 9.43 | 24.10 ± 13.16 | 28.19 ± 13.38 | 33.34 ± 24.90 | <0.001 |
| Median (Q1, Q3) | 15.00 (11.25,<br>19.40) | 22.60 (15.00,<br>30.60) | 29.30 (24.2, 36.7) | 22.0 (14.4, 46.9) |  |
| **HbA1c [%]** |  |  |  |  |  |
| Mean ± SD | 5.42 ± 0.26 | 5.43 ± 0.27 | 5.35 ± 0.29 | 6.43 ± 0.68 | <0.001 |
| Median (Q1, Q3) | 5.50 (5.30, 5.60) | 5.40 (5.20, 5.60) | 5.40 (5.15, 5.60) | 6.35 (5.85, 6.85) |  |
| **Total cholesterol** |  |  |  |  |  |
| **[mmol/l]** |  |  |  |  |  |
| Mean ± SD | 4.17 ± 0.63 | 3.82 ± 0.66 | 4.17 ± 0.77 | 4.00 ± 0.80 | 0.042 |
| Median (Q1, Q3) | 4.09 (3.74, 4.55) | 3.82 (3.36, 4.37) | 3.91 (3.57, 4.51) | 4.03 (3.50, 4.55) |  |
| **HDL-cholesterol** |  |  |  |  |  |
| **[mmol/l]** |  |  |  |  |  |
| Mean ± SD | 1.27 ± 0.34 | 1.17 ± 0.27 | 1.12 ± 0.22 | 1.07 ± 0.28 | 0.028 |
| Median (Q1, Q3) | 1.27 (1.0, 1.44) | 1.15 (0.73, 1.29) | 1.04 (0.95, 1.17) | 1.06 (0.83, 1.23) |  |
| **Triglyceride** |  |  |  |  |  |
| **[mmol/l]** |  |  |  |  |  |
| Mean ± SD | 1.10 ± 0.47 | 1.14 ± 0.58 | 1.41 ± 0.80 | 1.28 ± 0.60 | 0.057 |
| Median (Q1, Q3) | 0.92 (0.72, 1.4) | 1.04 (0.72, 1.36) | 1.04 (0.73, 1.34) | 1.16 (0.79, 1.63) |  |
| **Uric Acid [μmol/l]** |  |  |  |  |  |
| Mean ± SD | 301.7 ± 67.2 | 348.5 ± 85.4 | 371.9 ± 91.0 | 333.5 ± 46.7 | <0.001 |
| Median (Q1, Q3) | 304.4 (253.1, 351.2) | 317.6 (265.6, 348.6) | 370.3 (301.5, 429.1) | 330.8 (301.0, 355.8) |  |
| **ASAT [μkat/l]** |  |  |  |  |  |
| Mean ± SD | 0.51 ± 0.12 | 0.45 ± 0.11 | 0.43 ± 0.13 | 0.44 ± 0.18 | 0.013 |
| Median (Q1, Q3) | 0.49 (0.42, 0.59) | 0.43 (0.36, 0.48) | 0.40 (0.32, 0.51) | 0.37 (0.33, 0.48) |  |
| **ALAT [μkat/l]** |  |  |  |  |  |
| Mean ± SD | 0.48 ± 0.23 | 0.47 ± 0.22 | 0.53 ± 0.34 | 0.59 ± 0.36 | 0.433 |
| Median (Q1, Q3) | 0.40 (0.33, 0.45) | 0.41 (0.31, 0.52) | 0.42 (0.29, 0.64) | 0.46 (0.36, 0.72) |  |

Fasting glucose and HbA1c levels were significantly elevated in the T2DM group compared to non-diabetic participants (both p < 0.001). Fasting insulin concentrations steadily increased across puberty (p < 0.001).

Lipid parameters showed a significant decline in HDL cholesterol during puberty (p = 0.028), while triglyceride levels tended to increase (p = 0.057). Uric acid levels increased significantly with pubertal progression (p < 0.001). AST and ALT liver enzymes displayed minimal variations between groups with AST reaching statistical significance (p = 0.013) (Table 1).

### 3.1 Mitochondrial respiration and ATP production

Mitochondrial respiratory parameters were compared between the pubertal groups and between participants with obesity and IS/IR as well as participants with manifest T2DM. (Table 2). Coupling efficiency was significantly decreased in T2DM participants. BHI values compared across pubertal stages and insulin sensitivity status showed significantly higher BHI for postpubertal and insulin-resistant (IR) group compared to participants with T2DM (p = 0.0252). No significant differences were observed between IS and IR subgroups in all pubertal stages stages (Figure 1). Absolute values in ATP production from ATP Rate Assay did not show differences between the groups (Table 2). Relative numbers of mito ATP and glyco ATP showed significantly lower mito ATP and higher glyco ATP levels in T2DM participants compared to those without manifested T2DM (Figure 2).

**Figure 1:**
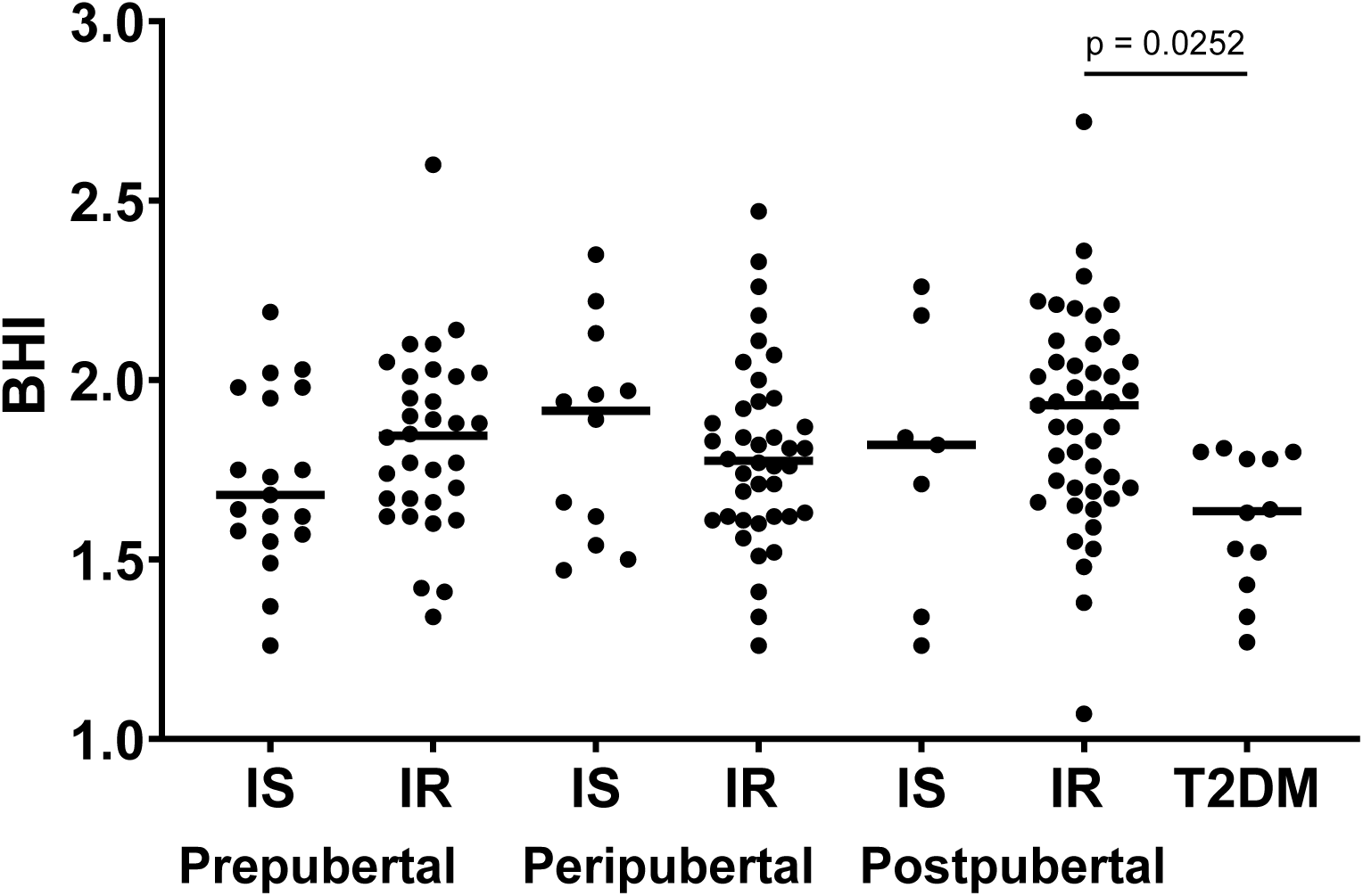
Bioenergetic Health Index (BHI) calculated from Mito Stress Test depending on pubertal stage, degree of insulin resistance and T2DM. Significantly lower BHI in T2DM. No significant differences between insulin sensitive (IS) and insulin resistant (IR) groups. No differences between pubertal stages. Significantly lower BHI in T2DM than postpubertal IR.

**Figure 2:**
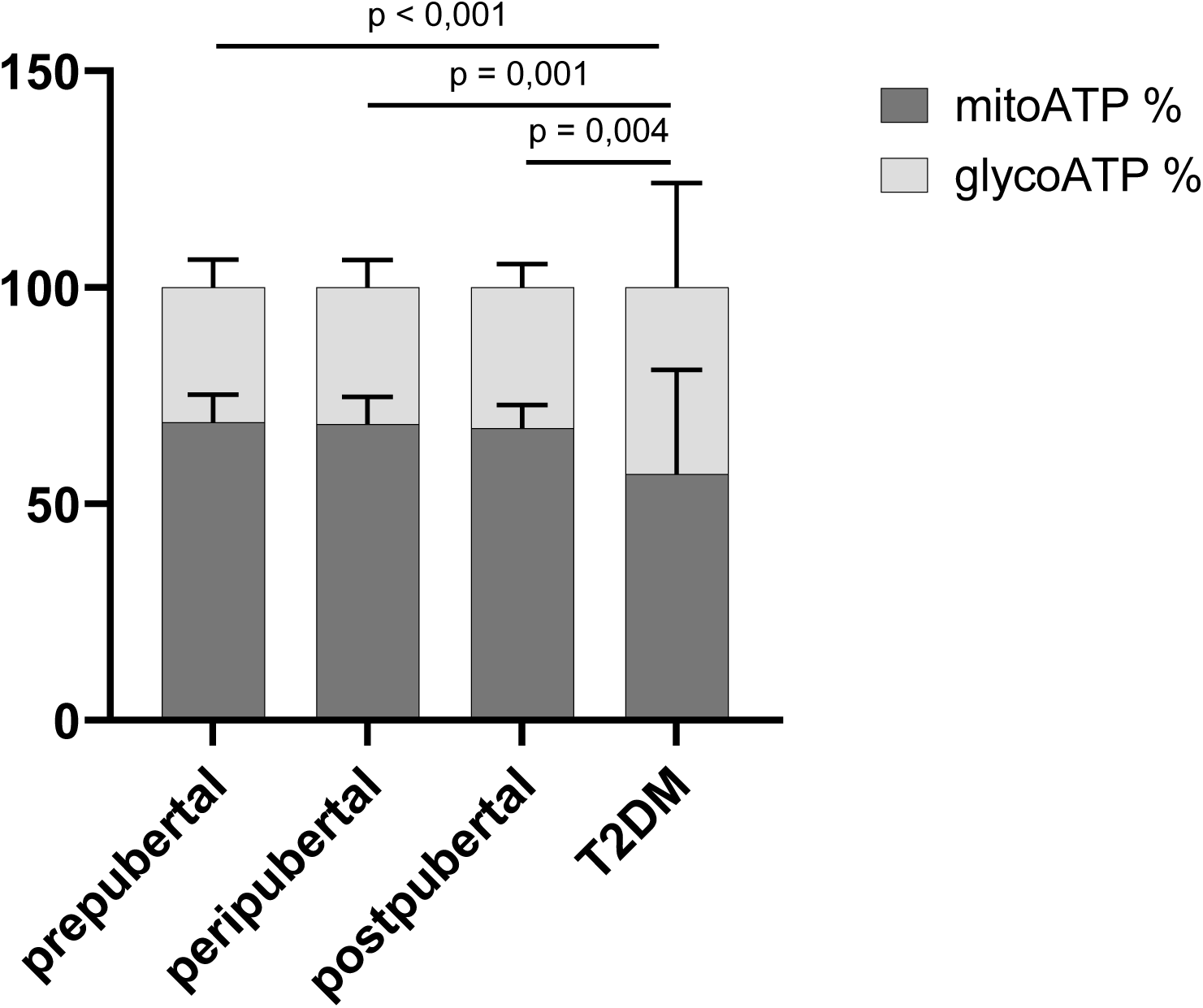
Ratio of mito ATP to glyco ATP from ATP Rate Assay depending on pubertal stage and T2DM. Significantly higher glyco ATP and lower mito ATP in T2DM compared to all pubertal stages without T2DM.

**Table 2:**
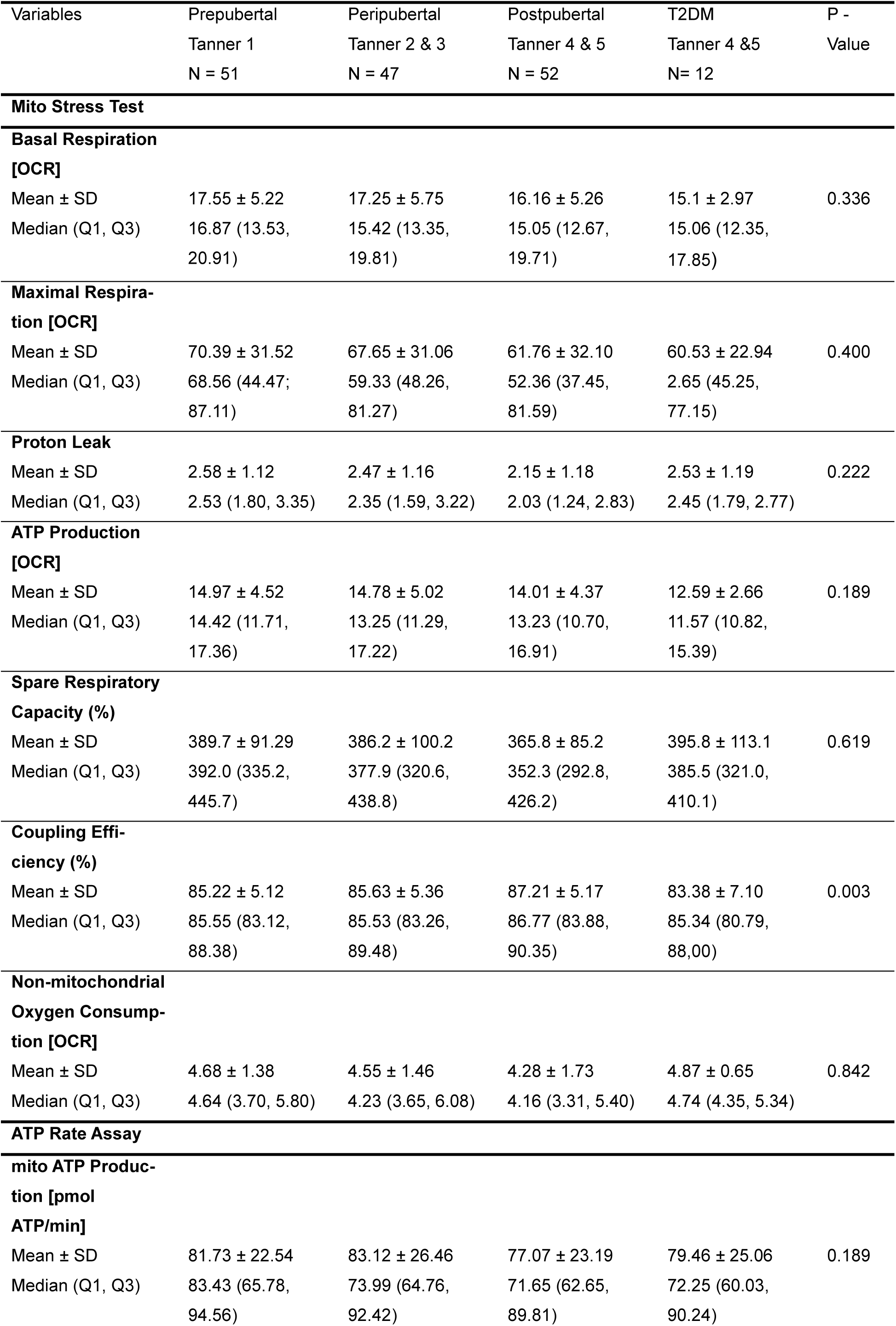
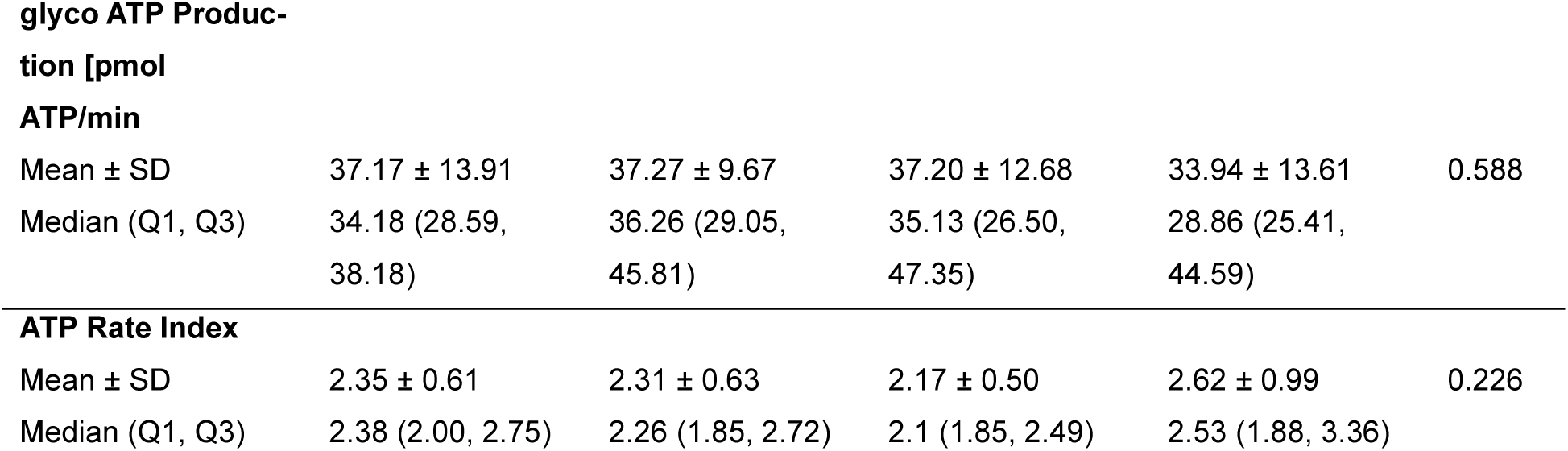
Mitochondrial respiration rates from Mito Stress Test and ATP Production rates from ATP Rate Assay.

**Table 3:** Spearman’s Correlation Analysis between mitochondrial respiration / ATP production parameters and IR / T2DM related parameters (n = 150).

|  | Mito Stress Test |  |  |  |  |  | ATP Rate Assay |  |  |  |  |
| --- | --- | --- | --- | --- | --- | --- | --- | --- | --- | --- | --- |
|  | Basal Re-<br>spiration | ATP-Pro-<br>duction re-<br>lated OCR | Maximal<br>Respiration | Proton<br>Leak | Spare<br>Respiratory<br>Capacity<br>(%) | Coupling<br>Efficiency<br>(%) | glycoATP<br>Production | mitoATP<br>Production | ATP Rate<br>Index | glycoATP<br>(%) | mitoATP<br>(%) |
| **BMI-SDS** |  |  |  |  |  |  |  |  |  |  |  |
| **ρ** | 0,127 | 0,109 | ,200* | 0,071 | ,210* | -0,057 | ,223** | 0,112 | -0,096 | 0,093 | -0,093 |
| **p** | 0,124 | 0,186 | 0,015 | 0,389 | 0,011 | 0,493 | 0,007 | 0,171 | 0,255 | 0,267 | 0,267 |
| **Age** |  |  |  |  |  |  |  |  |  |  |  |
| **ρ** | -0,122 | -0,102 | -0,111 | -0,122 | -0,020 | ,162* | 0,000 | -0,112 | -0,132 | 0,131 | -0,131 |
| **p** | 0,140 | 0,215 | 0,182 | 0,138 | 0,809 | 0,048 | 0,999 | 0,171 | 0,116 | 0,119 | 0,119 |
| **WHtR** |  |  |  |  |  |  |  |  |  |  |  |
| **ρ** | 0,092 | 0,092 | ,184* | 0,023 | ,216** | 0,021 | 0,145 | 0,043 | -0,042 | 0,043 | -0,043 |
| **p** | 0,266 | 0,265 | 0,026 | 0,778 | 0,008 | 0,801 | 0,084 | 0,600 | 0,614 | 0,614 | 0,614 |
| **fasting Glucose** |  |  |  |  |  |  |  |  |  |  |  |
| **ρ** | 0,120 | 0,131 | 0,135 | 0,070 | 0,077 | 0,029 | 0,040 | 0,113 | 0,102 | -0,097 | 0,097 |
| **p** | 0,147 | 0,109 | 0,102 | 0,394 | 0,355 | 0,730 | 0,632 | 0,170 | 0,224 | 0,249 | 0,249 |
| **fasting insulin** |  |  |  |  |  |  |  |  |  |  |  |
| **ρ** | 0,035 | 0,023 | 0,071 | -0,049 | 0,055 | 0,084 | ,197* | -0,020 | -,198* | ,193* | -,193* |
| **p** | 0,671 | 0,783 | 0,392 | 0,557 | 0,505 | 0,306 | 0,018 | 0,811 | 0,018 | 0,021 | 0,021 |
| **HOMA-IR** |  |  |  |  |  |  |  |  |  |  |  |
| **ρ** | 0,040 | 0,030 | 0,080 | -0,049 | 0,064 | 0,093 | ,186* | -0,017 | -,175* | ,172* | -,172* |
| **p** | 0,628 | 0,719 | 0,338 | 0,555 | 0,443 | 0,259 | 0,026 | 0,838 | 0,036 | 0,040 | 0,040 |
| **HbA1c** |  |  |  |  |  |  |  |  |  |  |  |
| **ρ** | 0,027 | -0,001 | -0,021 | 0,126 | -0,053 | -0,142 | 0,084 | 0,057 | -0,046 | 0,057 | -0,057 |
| **p** | 0,747 | 0,991 | 0,800 | 0,126 | 0,523 | 0,085 | 0,318 | 0,488 | 0,585 | 0,497 | 0,497 |

Mitochondrial respiration parameter in postpubertal participants stratified in IS (n = 7), IR (n = 45) and T2DM (n = 12) confirm previous results in lower BHI (p = 0.0099) and Coupling efficiency (p < 0.001) in T2DM than IR. The three groups showed no significant variations in their basal respiration levels and OCR related to ATP production, proton leak, nonmitochondrial respiration and spare respiratory capacity (Figure 3).

**Figure 3:**
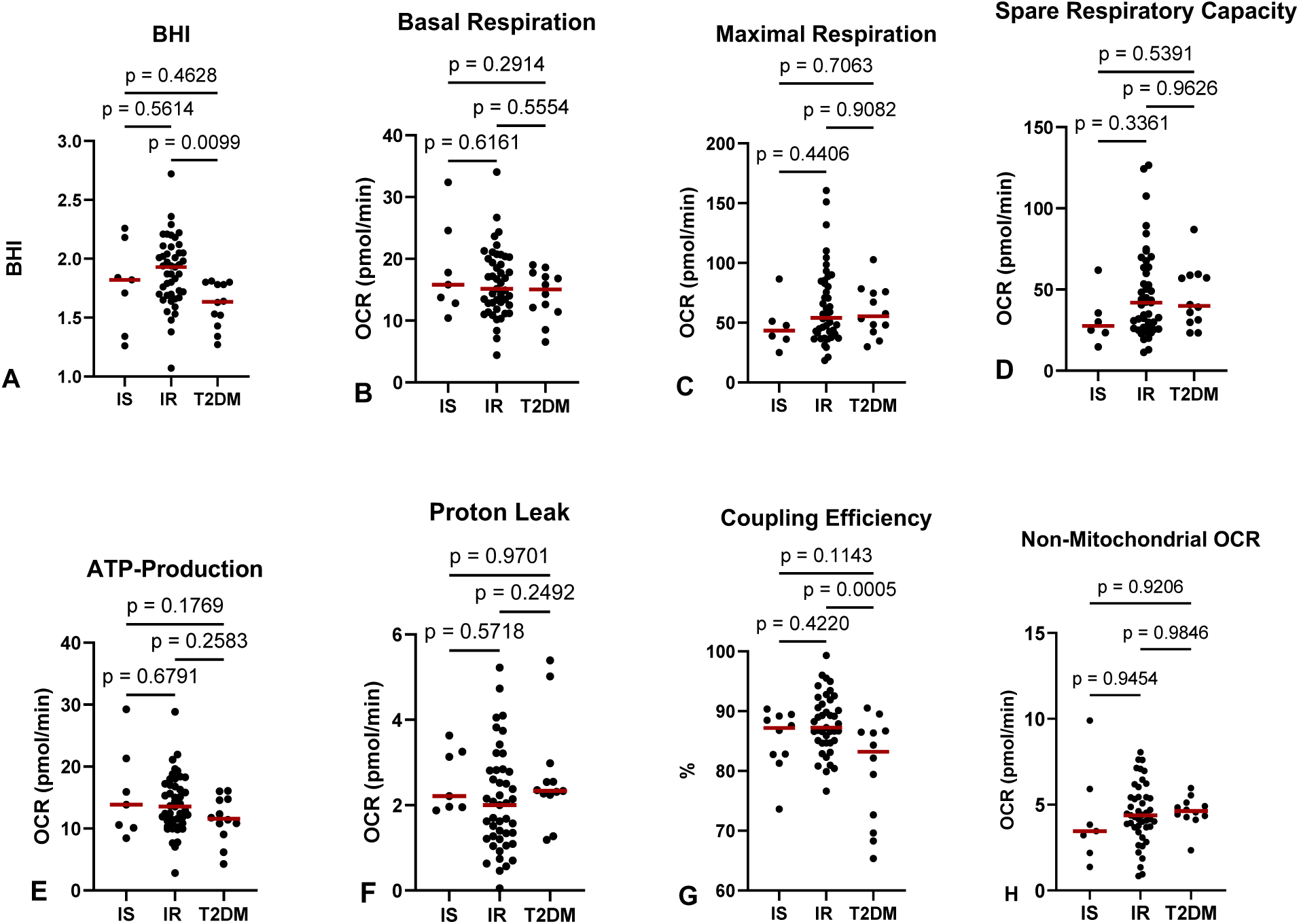
Mitochondrial function in postpubertal, insulin-resistant participants with (IR) and without insulin resistance (IS) or Type 2 Diabetes Mellitus (T2DM). A: Bioenergetic Health Index (BHI) was significantly lower in T2DM than in IR. B: No difference in Basal Respiration Oxygen Consumption Rate (OCR). C: No difference in OCR during maximal respiration. D: No difference in spare respiratory capacity. E: No difference in ATP production-related OCR. F: No difference in proton leak. G: Significantly lower coupling efficiency in T2DM than IR. H: No difference in non-mitochondrial OCR.

## 4. Discussion

This study has investigated mitochondrial respiration in fresh PBMCs in children and adolescents with obesity and varying degrees of insulin resistance compared to adolescents with obesity and manifest T2DM. Gender, different pubertal stages and different stages of IR were considered. We aimed to a) investigate whether metabolic health impairs with increasing stages of IR or manifest T2DM on children with obesity and b) to assess the suitability of PBMCs as a minimally invasive biomarker for metabolic health in pediatric populations.

To the best of our knowledge, this is the first study that has investigated markers of mitochondrial function in PBMC cells in pre-, periand postpubertal children with obesity and IR. The findings provide several important insights into the utility of mitochondrial bioenergetic profiling in the context of pediatric obesity and metabolic disease.

Our data reveal that mitochondrial respiration in fresh PBMCs is sensitive to the degree of metabolic impairment in childhood obesity. The findings for the T2DM group show lower coupling efficiency, which reflects how efficiently oxygen consumption is used to produce ATP. This indicates a reduced ability to handle metabolic stress, suggesting mitochondrial alterations, impaired energy production, and reduced metabolic flexibility. Possible biological implications could be oxidative stress, cellular senescence or chronic metabolic disease.

Although basal respiration and ATP production related OCR did not significantly differ between groups, the BHI was significantly lower in individuals with T2DM compared to those with insulin resistance and same pubertal stage (i.e., postpubertal). This suggests that the BHI, a composite index reflecting multiple aspects of mitochondrial respiration, may amplify small differences in parameters, serving early indicator of mitochondrial alterations than isolated respiratory parameters [30, 31]. Simultaneously, the BHI does not show in which process of mitochondrial respiration the alterations are present. These results align with earlier research demonstrating associations between PBMC bioenergetics and metabolic status in adults and support the potential of BHI as a biomarker in pediatric metabolic health assessments [15, 19].

Furthermore, the observed decrease in BHI in participants with T2DM supports the growing evidence that mitochondrial changes are not merely a consequence of long-standing metabolic disease but may play a central role in its early pathogenesis [33]. These alterations in mitochondrial performance likely reflect increased oxidative stress, diminished ATP production efficiency, and altered mitochondrial dynamics, all of which have been implicated in insulin resistance and the progression of metabolic syndrome development [12, 34].

Despite clear metabolic stratification based on HOMA-IR percentiles, the mitochondrial respiration in insulin resistant participants does not shift towards the mitochondrial respiration in T2DM participants. Interestingly, the data from postpubertal, IR children indicate a trend towards higher mitochondrial respiration rates. This finding might reflect mitochondrial adaptation or compensation processes of chronic overnutrition or chronic elevated blood glucose levels. Relative ATP production through glycolysis is weakly correlated with increased fasting insulin levels and HbA1c but not with fasting glucose. However, the ATP production from glycolysis did only increase in T2DM participants and not yet in IR children. The elevated mitochondrial respiration in PBMCs, which also includes IR participants’ immune cells, may also be attributed to increased systemic inflammation. Previous studies showed a strong correlation between HOMA-IR and inflammatory markers such as interleukin-6 (IL-6) or C-reactive protein (CRP) [35]. However, this hypothesis does not account for the observed alteration in mitochondrial respiration following the onset of T2DM, despite the continued progression in inflammation [36]. Böhm et al. showed similar findings in adipocytes in adults: subjects with insulin resistance had elevated respiration rates, compared to participants with insulin-sensitive obesity [37, 13].

Carvalho et al. have also investigated mitochondrial respiration in PBMC cells from children with overweight or obesity. The authors showed – among other results - that PBMC respiration positively correlated with BMIz, HOMA-IR and fasting glucose and insulin, but was negatively correlated with inflammatory cytokines in prepubertal children. The authors concluded that PBMCs from young children with overweight/obesity may already exhibit adaptations to the metabolic stressors associated with insulin resistance and that PBMC metabolism correlates well with whole-body metabolism [38]. However, in contrast to our study, they have only investigated prepubertal children (ages 5-10 years). Thus, we confirm and extend these findings by showing that parameters of metabolic health impair with increased insulin resistance and show significantly disturbance in states of manifest T2DM in youths with obesity.

Similar results in adults were recently reported by Pinho et al. and Barbosa et al. [13, 39]. This study evaluated and compared bioenergetics and energy substrate preference by omental and subcutaneous adipose tissue from 40 adult subjects with obesity at distinct metabolic stages. In accordance with our results, the authors could show that oxidative phosphorylation capacity of adipose tissue differs through the progression of metabolic disease and that subjects with obesity and diabetes had the lowest capacity in both, subcutaneous and omental adipose tissues [39].

Thus, our findings confirm and extend the results of both studies from Carvalho et al. and Barbosa et al. by showing that there might already be several mitochondrial adaptation processes to chronic overnutrition in pre-, peri-, and postpubertal children with obesity and that these compensatory mechanisms fail and collapses as soon as progression to T2DM has occurred.

### 4.1 Strengths and limitations

Our study cohort is an anthropometrically, clinically, and metabolically well-defined group of children and adolescents with obesity, spanning all pubertal stages from Tanner stages 1 (prepubertal) to Tanner stages 5 (postpubertal). A total of 162 participants were included, representing a markedly higher number of participants compared to previously published studies. The categorization into subgroups based on age adjusted insulin resistance according to HOMA-IR percentiles or T2DM, respectively, provides a clearer picture of the relationship between mitochondrial alteration and metabolic impairment, while also minimizing confounders such as puberty and age. We were – to the best of our knowledge – also able for the first time to include a well definded group of adolescent patients with T2DM in our analyses. Another strength of this study lies in using fresh PBMCs, a minimally invasive and easily accessible surrogate tissue, for mitochondrial function assessment in a pediatric population. Prior studies have demonstrated that PBMCs can reflect metabolic activity in less accessible tissues such as muscle and liver [14, 16]. The current research results demonstrate their practical value for pediatric care, especially for identifying metabolic disturbances at an early stage. We have applied the Bioenergetic Health Index (BHI), a well-defined and validated marker for metabolic health, in pediatric patients with different levels of metabolic impairment We show for the first time and regard this as a major strengths of our study, that its application in children and adolescents with obesity and T2DM may represent a new diagnostic tool that provides essential knowledge about mitochondrial bioenergetics in early metabolic disease progression. The use of minimally invasive sampling methods (PBMCs from venous blood) makes this approach ethically and logistically suitable for pediatric research, supporting its potential translation into routine clinical practice for early detection and risk assessment.

The main limitation of this study is the sample size of the subgroup of participants with manifest T2DM, as T2DM is still regarded as a rare disease in the pediatric population and historically been considered a disease of adulthood. Second, while we controlled for key confounding variables, such as age and BMI-SDS, other factors such as physical activity levels or dietary intake, known to influence mitochondrial function and insulin sensitivity, were not systematically assessed. Additionally, although PBMCs offer a convenient and minimally invasive model, they may not fully capture the tissue-specific mitochondrial adaptations occurring in metabolically active tissues such as liver, skeletal muscle, or adipose tissue. Moreover, PBMCs are representing a heterogeneous mixture of immune cells, including lymphocytes and monocytes, each with potentially distinct mitochondrial characteristics and bioenergetic profiles. Without cell-type-specific analysis, it remains unclear to what extent observed differences in mitochondrial function are driven by true mitochondrial dysfunction versus shifts in cellular composition.

## 5. Conclusion

PBMC-based mitochondrial bioenergetic profiling may serve as a sensitive biomarker for early metabolic impairment in children and adolescents with obesity and several degrees of insulin resistance. Bioenergetics in postpubertal participants with obesity and T2DM is significantly lower compared to postpubertal participants with obesity and insulin resistance. Further studies are needed to validate its clinical utility, and immunophenotyping or single-cell approaches should be included to better account for this heterogeneity and to evaluate whether specific subpopulations contribute disproportionately to the observed mitochondrial alterations.

## Supporting information

Graphical Abstract

## Acknowledgments

We would like to express our sincere gratitude to all children and adolescents and their parents as well as caregivers who have participated in this study. We thank Lorenz Greifoner, Dinet Ahmed and Leah Tomerius for assisting during the study visits and data organization as well as Ines Volkmer for supporting during laboratory work. We would also like to thank Prof. Andreas Wienke for counselling for statistical analysis.

## CRediT authorship contribution statement

Writingoriginal draft: MR, SWB. Writing – Review and editing: all authors. Visualization, Formal analysis, Investigation, Resources: MS, SWB, SAR, SK, AS, EC. Methodology: MS, SWB, SAR, EC, HZ, AS, PJO. Conceptualization and Data curation: MR, EC, SK, PJO, SWB. Funding acquisition: PJO, EC, SWB.

## Institutional Review Board Statement

The study was conducted according to the guidelines of the Declaration of Helsinki, and approved by the Institutional Review Board (or Ethics Committee) of the Ethics Committee of the University Medicine Halle (Ethic’s vote of 24^th^ of June 2024, # 2023-238). Participants and their parents or legal guardians were informed both in writing and verbally about the study procedures and data analysis and provided written consent. The study was performed according to the STROBE criteria.

## Data Availability Statement

The authors confirm that clinical, anthropometrical and biochemical parameters were collected in our center from both study cohorts. All data generated or analyzed during this study are included in this article. However, due to protection of patient privacy and the specifications in the patient/parent consent form, we may not share patient level data with researchers outside University Medicine Halle. Further enquiries can be directed to the corresponding author.

