## Supplementary material for "Decreased Mitochondrial Respiration in Peripheral Mononuclear Blood Cells in Children and Adolescents with Obesity and Type 2 Diabetes Mellitus": Graphical Abstract

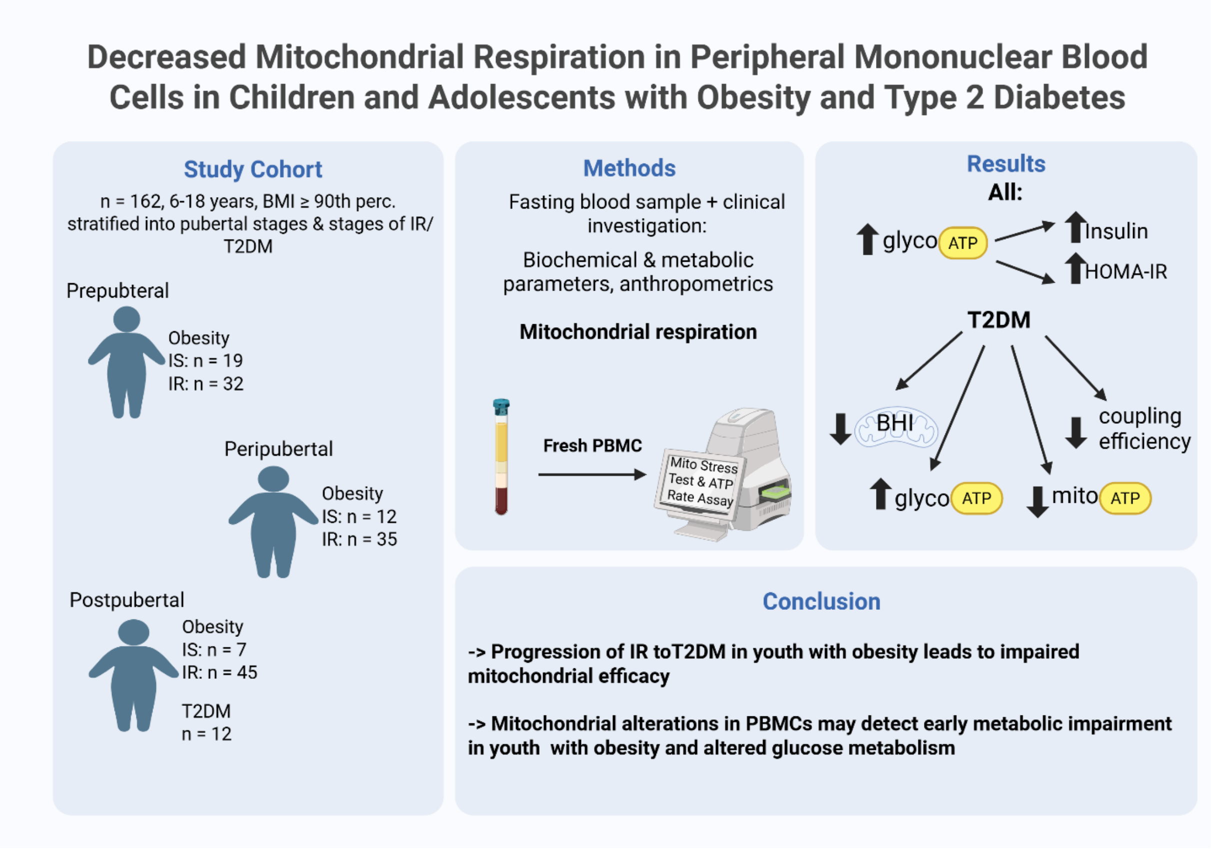


Created with BioRender.com
